# Sex-biased Genetic Risk Loci and Causal Brain Proteins in Parkinson’s Disease

**DOI:** 10.64898/2026.06.23.26356345

**Authors:** Noah Cook, Youjie Zeng, Tianyi Fu, Chenyu Yang, Sathesh K. Sivasankaran, Phuc Nguyen, FinnGen, Aliza P. Wingo, Thomas S. Wingo, Jia Nee Foo, Albert A. Davis, Laura Ibanez, Carlos Cruchaga, Michael E. Belloy

**Author notes:** **Corresponding Author** Michael E. Belloy, Department of Neurology, NeuroGenomics and Informatics Center (NGI), Washington University in Saint Louis (WashU), 4444 Forest Park Ave, St. Louis, MO 63108, USA. Shared first authors (Noah Cook, Youjie Zeng, Tianyi Fu, Chenyu Yang, Michael E. Belloy).

## Abstract

Parkinson’s disease (PD) exhibits pronounced sex differences, yet the underlying genetic and molecular mechanisms remain poorly understood. We performed the largest-to-date meta-analysis of sex-stratified genome-wide association studies of PD followed by brain proteogenomics-based causal inference analyses. We nominated 10 candidate proteins that appear important to sex-biased PD risk, of which 2 female-biased, GALC and PSMG1, and 3 male-biased, ACTR1B, WDR41, and CD151, were most robustly prioritized. Together, our findings provide evidence for genetic sex differences in PD, prioritizing sex-biased proteins implicated in lysosomal regulation, neuroinflammation, lipid biology, and other PD-relevant mechanisms, and highlighting potential sex-informed therapeutic opportunities.

## Introduction

Parkinson’s disease remains a debilitating disease without an effective disease-modifying therapy^1^. New discoveries are anticipated by capitalizing on the pronounced sex dimorphism in PD. Men account for two-thirds of incident PD cases and differ from women in disease age-at-onset, symptom patterns, progression, and therapy response^2–4^. While this may relate to hormonal, environmental, and lifestyle factors, little is known about causal genetic and molecular mechanisms that could underlie this dimorphism. The first sex-stratified PD GWAS did not identify sex differences for genome-wide significant variants^5^. However, subsequent work integrating non-sex-stratified GWAS of PD with GWAS of age-at-menopause and age-at-menarche identified pleiotropic loci^6^. This marked candidate PD risk variants and genes that may be modulated by sex and highlighted a role for immune response pathways in PD in males^6^. Yet, whether these genetic signals are causally shared across sex hormone traits, sex-biased PD risk, and the implicated genes remains unverified, leaving key questions unresolved. Wingo *et al*. created a brain proteogenomics resource of sex-stratified variant-to-protein associations (protein quantitative trait loci [pQTLs]), which they integrated with the aforementioned sex-stratified PD GWAS^7^. This approach, exploring genetically regulated proteins for overlap with the genetics of a given disease is termed a proteome-wide association study (PWAS) and, together with complimentary methods, enables inference of disease causal proteins^7–9^. Although they nominated four sex-biased PD proteins, the analyses were limited by the power of the underlying sex-stratified PD GWAS and were restricted to using sex-stratified pQTLs for gene nomination. Prior studies have focused on sex-biased variant-to-gene associations to elucidate sex differences in human traits, but few variant-to-gene associations display sex-biases^7,10^. Our recent work in Alzheimer’s disease showed that leveraging non-sex-stratified pQTL data amplified insights into sex-biased Alzheimer’s disease genetic signals^9^, highlighting the opportunity for similar analyses in PD. Here, we overcome these gaps by expanding the sample size of sex-stratified PD GWAS and broadening proteogenomics causal inference analyses to implicate candidate sex-biased PD proteins.

## Methods

Participants or their caregivers provided written informed consent in the original studies. The study protocol was granted an exemption by the Washington University Institutional Review Board because the analyses were carried out on “de-identified, off-the-shelf” data; therefore, additional informed consent was not required. The FinnGen ethics statement is available in the supplement (Nr HUS/990/2017).

Sex-stratified PD GWAS were available from Blauwendraat *et al*.^5^, which included GWAS from the International Parkinson’s Disease Genomics Consortium (IPDGC) in clinically ascertained subjects, and GWAS from UK Biobank, conducted in ICD-defined cases versus controls and proxy-cases (mother or father affected with PD) versus controls. We newly included sex-stratified PD GWAS of ICD-defined cases versus controls in FinnGen (cf. Supplement)^11^. All data pertained to subjects of European ancestry. GWAS per sex were combined using fixed effects meta-analyses. Independent GWAS signals were determined through LD clumping (window=2Mb, R^2^<0.01) of signals passing genome-wide significance (P<5×10^-8^). Lead variants were assessed for sex-heterogeneity using 1) heterogeneity tests on meta-analyzed effect estimates, Z=(Beta_Men_-Beta_Women_)/√(SE_Men_^2^+SE_Women_^2^), and 2) fixed effect inverse-variance weighted meta-analysis of the differences in male and female effect sizes per cohort, ΔBeta_i_=(ΔBeta_i,Men_-ΔBeta_i,Women_) and SE_i_=√(SE_i,Men_^2^+SE_i,Women_^2^) where i=cohort. Nominally significant signals on both tests, respectively referred to as P_Sex-Het_<0.05 and P_Sex-Het-Meta_<0.05, were considered sex-biased. Those passing additional FDR-correction for the number of independent genome-wide significant signals across either men or women were considered more robustly sex-biased. To assess power in respective sex-stratified GWAS, effective sample sizes were calculated as N_Eff_=4*v*(1-v)^12^, where v=N_Cases_/(N_Cases_+N_Controls_) and sample contributions from proxy-GWAS were divided by 4 to account for their reduced power^12^. This approach rescales a study’s sample size to that for an equivalently powered study with a balanced sample design.

Human brain proteomes were previously generated using isobaric tandem mass tag peptide labelling coupled with liquid chromatography-mass spectrometry (TMT-MS) from the dorsolateral prefrontal cortex of brain samples from European ancestry participants in the Religious Orders Study (ROS) and Memory Aging Project (MAP)^7^. Among 808 participants with proteomics data, 8,458 proteins passed quality control and were used for non-sex-stratified and sex-stratified pQTL analyses, as well as generation of proteogenomic weight files for PWAS (FUSION)^13^. For additional technical details, cf. supplement.

To identify candidate PD proteins with robust causal inference support, we required positive findings across three complimentary methods. First, we performed PWAS and identified proteins displaying P_FDR_<0.05. PWAS followed a primary discovery design in which GWAS and protein weights were sex-matched and a secondary design in which sex-specific GWAS were combined with non-sex-stratified protein weights^9^. Next, we performed genetic colocalization (COLOC) and summary-based Mendelian randomization (SMR) across pQTL and GWAS data (cf. supplement). Both methods aimed to corroborate that respective genetic signals causally influenced protein abundance, and in turn, PD risk. SMR analyses were complimented by HEIDI tests to reduce false positives. Positive support was considered for colocalization probability >0.7 and SMR-P_FDR_<0.05 with P_HEIDI_>0.05. All analyses followed standard approaches and significance criteria^7–9^.

Finally, causal inference was evaluated from two perspectives. First, sex-biased genome-wide significant PD signals were evaluated for evidence of causal proteins. Second, since PWAS leverage GWAS signals below genome-wide significance, we identified additional sex-biased PWAS discoveries by selecting proteins that displayed P_FDR_<0.05 in one sex and P>0.05 in the other sex, consistent with prior studies^7,9^. We then assessed additional COLOC/SMR support and whether underlying PD genetic signals displayed nominal or FDR-significance (corrected for all sex-biased PWAS protein discoveries) for P_Sex-Het_ and P_Sex-Het-Meta_ to prioritize top candidate sex-biased PD proteins.

## Results

Sex-stratified PD GWAS included 328,900 male and 386,146 female subjects and showed no genomic inflation (**Figure-1A, eFigure-1**). Effective sample sizes were however larger in males (N_Eff_=65,820) than in females (N_Eff_=42,497), marking larger power in the male-stratified analyses. Of 23 independent genome-wide significant signals across either female or male-stratified GWAS, all were identified in prior non-sex-stratified PD GWAS (**eTable-1**). The *GALC* locus (OR_Females_=1.10, P_Females_=2.35x10^-8^, OR_Males_=1.03, P_Males_=1.17x10^-1^, P_Sex-Het_=1.28x10^-3^, P_Sex-Het-Meta_=1.29x10^-3^) showed a female-biased association passing FDR-significance on both sex-heterogeneity tests, while *SETD1A* (OR_Females_=0.89, P_Females_=6.17x10^-12^, OR_Males_=0.94, P_Males_=1.05x10^-5^, P_Sex-Het_=1.78x10^-2^, P_Sex-Het-Meta_=1.64x10^-2^), and *BST1* (OR_Females_=0.94, P_Females_=2.40x10^-4^, OR_Males_=0.90, P_Males_=1.20x10^-13^, P_Sex-Het_=4.34x10^-2^, P_Sex-Het-Meta_=3.57x10^-2^), respectively, showed female and male-biased associations passing nominal (not FDR) significance on both sex-heterogeneity tests (**Figure-1, eFigure-2**). Causal protein inference nominated GALC as the candidate PD gene at its respective locus (**Figure-2**).

**Figure 1.**
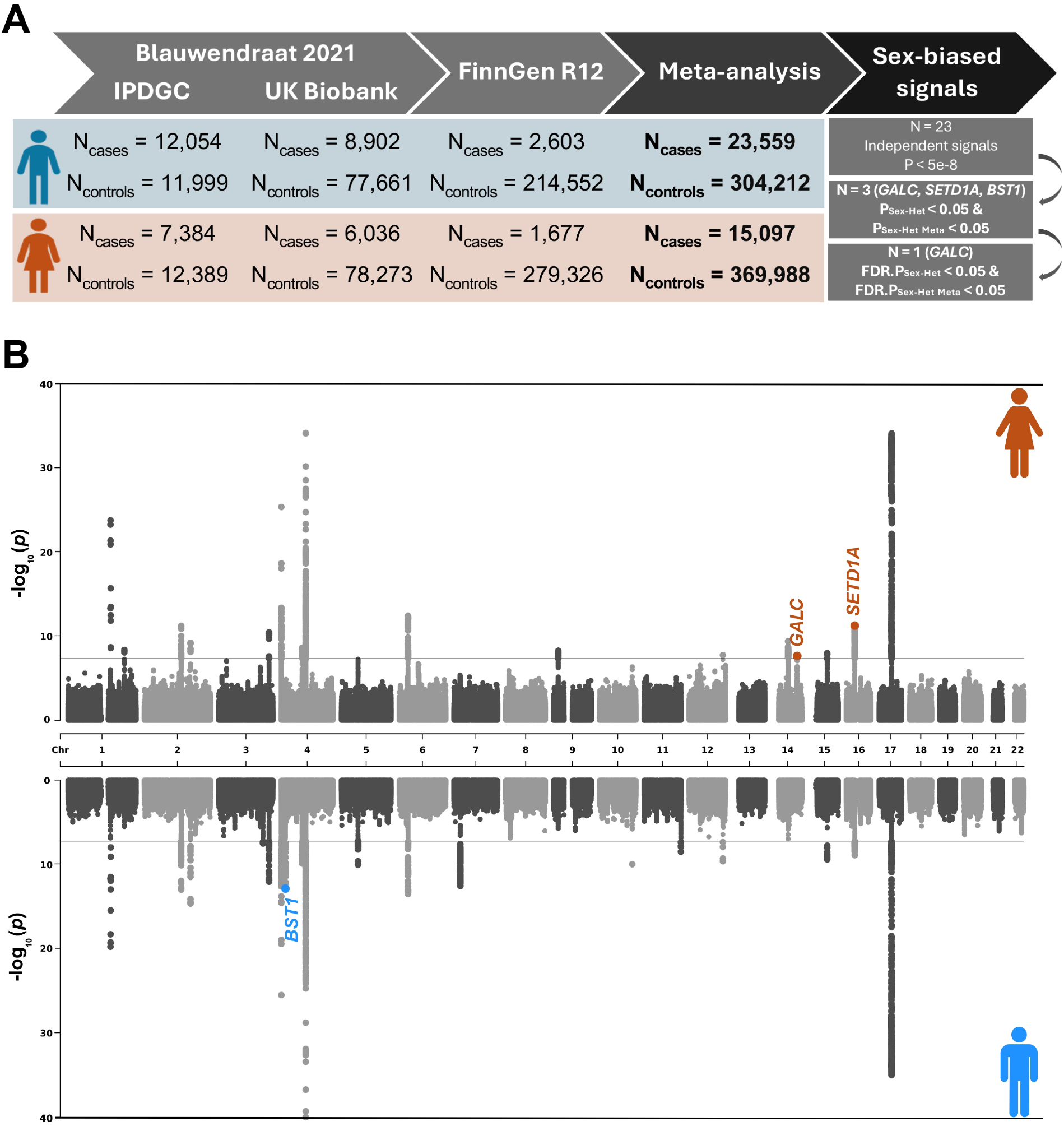
Sex-stratified Parkinson’s disease GWAS. **A)** Schematic overview of included cohorts, case-control counts, number of independent sex-stratified signals, and number of sex-stratified signals passing sex heterogeneity significance criteria. **B)** Miami plot displayed female (top) and male (bottom) stratified GWAS results. Lead variant at two sex-biased loci are labelled with their nearest gene.

Sex-stratified PWAS further identified 68 unique candidate proteins at P_FDR_<0.05 (37 male-only, 15 female-only, 16 shared), including 41 within known PD risk loci (**eTables-2-6, eFigures-3-4**). Of these, 28 (23 male-only) passed PWAS sex-heterogeneity filters, of which 9 showed convergent support from COLOC/SMR and mapped to subthreshold (P<5e-8) PD GWAS signals (**Figure-2;** Extended results and information regarding COLOC/SMR is presented in **eTables-7-8**). Out of these 9 proteins, 4 passed FDR and 1 (PTPN1) nominal (not FDR) significance on both sex-heterogeneity tests of the underlying PD GWAS signals. Altogether across GWAS and PWAS, five proteins–GALC, PSMG1, ACTR1B, WDR41, and CD151– were most robustly prioritized, passing FDR-correction on all sex heterogeneity tests. Locus novelty relative to prior PD GWAS and genetic effect estimates, together with sex-heterogeneity P-values, are provided in **Figures-2-3**.

**Figure 2.**
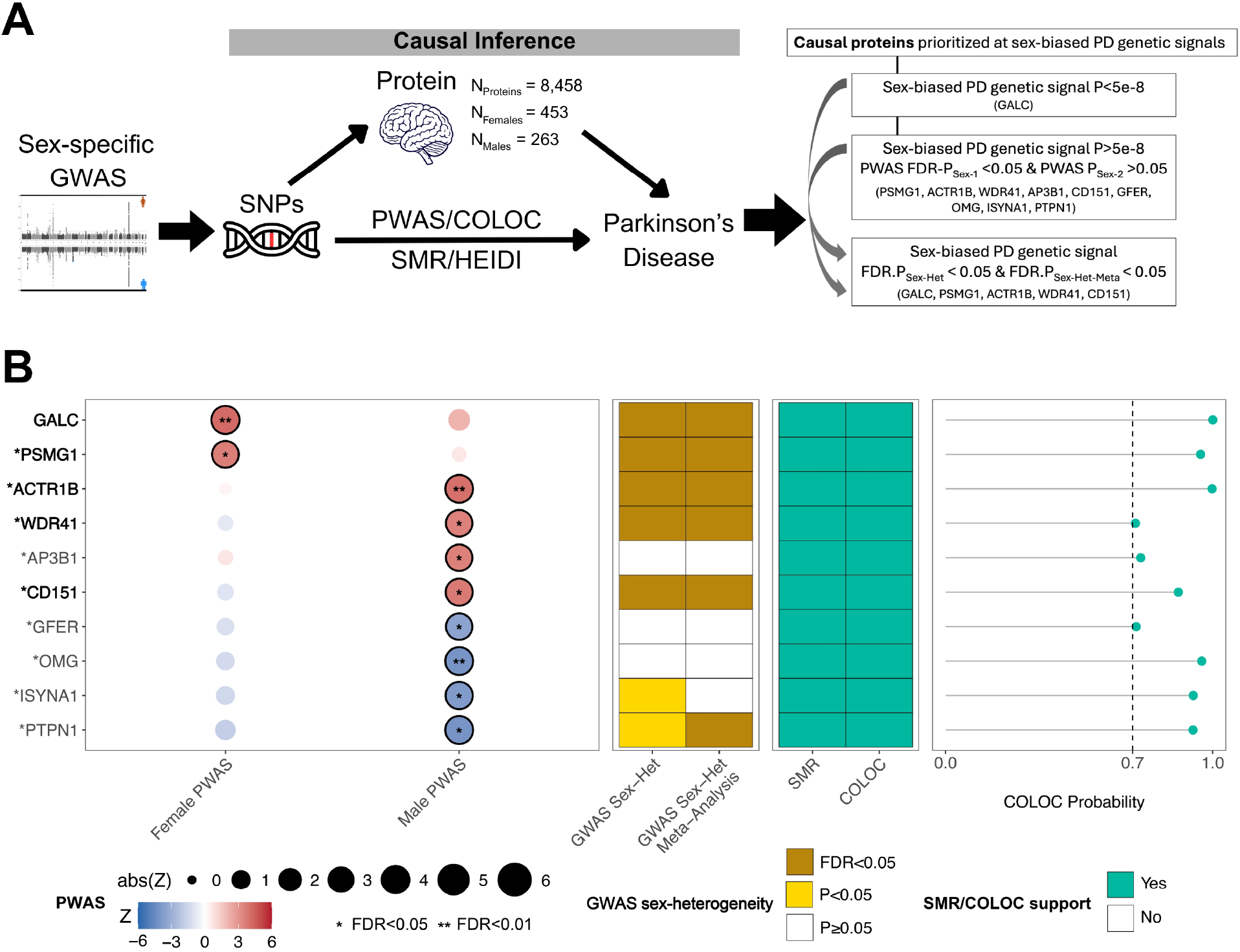
Inference of sex-biased candidate PD causal proteins. **A)** Schematic overview causal protein inference analyses (left) and summary (right) of top prioritized proteins supported by all three causal inference approaches. **B)** Overview matrix of findings for 10 prioritized proteins, including sex-stratified PWAS findings (left), sex-heterogeneity summaries of underlying GWAS signals (middle), and support from SMR and COLOC analyses (right). Bolded gene names (left) indicate the 5 most robust proteins that additionally were FDR significant in GWAS sex heterogeneity analyses. Genes marked by * reflect those falling in novel PD risk loci.

**Figure 3.**
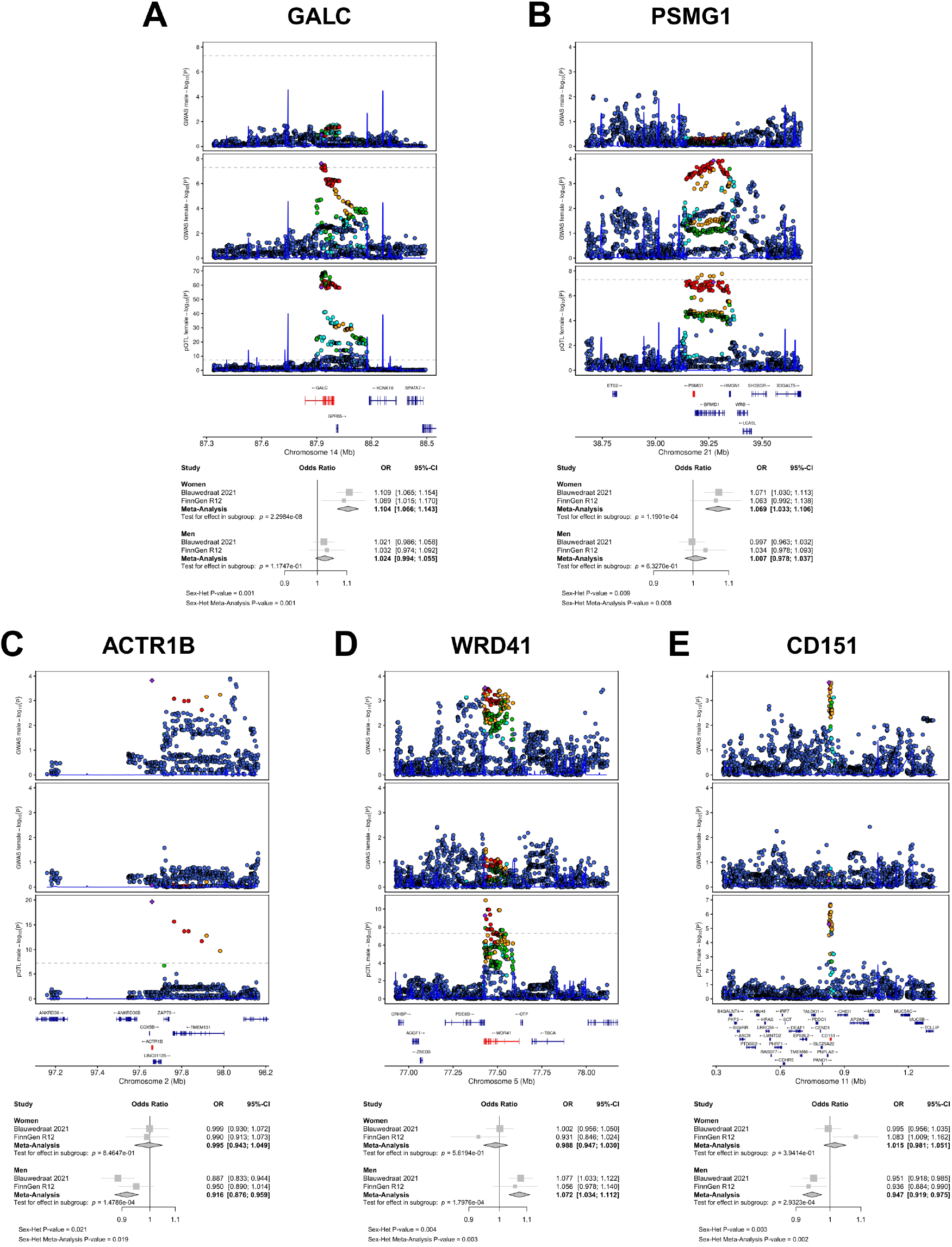
Locus zoom and forest plots for five top sex-biased PD candidate loci and proteins. Each set of panels show locus zoom plots for male PD GWAS (top), female PD GWAS (middle), and sex-matched colocalized pQTL data (bottom). The same tagging variant is indicated throughout. Dot color coding represents linkage with tagging variant (purple diamond): red - R2: 0.8-1.0, orange - R2:0.6-0.8, green - R2:0.4-0.6, cyan - R2:0.2-0.4, blue - R2:0.0-0.2. Forest plots indicate PD associations across cohorts and sexes for the tagging variant.

## Discussion

Our analyses prioritized five genetic risk loci and their candidate causal proteins that appear important to sex-biased PD risk. Despite sex-biased genetic associations at these loci, the genetic regulation of these proteins’ abundances was not sex-biased (**eTable-9**), which in part explains why they have not been nominated in prior studies that focused on proteins with evidence of sex-biased abundance^7^. This is important because these proteins can still impact mechanisms or pathways that contribute to sex-biased PD etiology, while evidence of sex-biased protein abundance may be lost in the postmortem proteomics samples due to the natural heterogeneity of PD, advanced age, or missing disease context (i.e. absence of PD cases). The *GALC* locus was the most robust sex-heterogeneous genome-wide significant signal we observed and additionally had GALC nominated as the candidate causal gene. Notably, *GALC* was already implicated to be of potential relevance in female-biased PD risk by prior efforts that mapped it to genetic loci that are pleiotropic across PD and estrogen-related traits^5^. *GALC* encodes galactosylceramidase, which impacts glycosphingolipid metabolism and prior work has suggested modulating galactosylceramidase activity as a therapeutic approach for PD^14^. PSMG1 (female-biased) and ACTR1B, WDR41 and CD151 (male-biased) represent discoveries at novel candidate PD risk loci that fall below conventional genome-wide significance, yet all display compelling links to known PD biology. PSMG1 (PAC1) functions in proteasome assembly and protein homeostasis^15^, ACTR1B is involved with intracellular transport and endolysosomal trafficking^16^, WDR41 regulates the autophagy-lysosomal pathway^17^, and CD151 has been linked to endolysosome function and T-cell activation^18,19^. Even PTPN1 (also known as PTP1B), which only passed nominal significance on sex-heterogeneity testing (**eFigure-5**), encodes protein tyrosine phosphatase 1B and has been linked to neuroinflammation and proposed as a therapeutic target for PD^20^. Mechanisms linking all proteins nominated in **Figure-2** are proposed in **eTable-10**.

### Limitations

The European ancestry of participants limits generalizability, underscoring the need for similar studies in diverse populations. While we identified more male-biased risk genes, this was likely mostly driven by the larger effective sample size (i.e. power) in the male GWAS. Larger female PD GWAS will thus be important to expand female-biased gene discoveries in the future. Overall, additional sex-stratified PD GWAS will be relevant to validate our novel protein discoveries at loci still below conventional genome-wide significance. Our proteogenomic samples originated from individuals without PD and were obtained from a single cortical brain region, so future proteogenomic data from PD-relevant cohorts and brain areas will be important to corroborate our findings and enable additional discoveries. It should finally be noted that our gene prioritization pertains to *in silico* approaches and may have missed other candidate causal genes co-regulated in other tissues or in other molecular layers (e.g. RNA expression).

## Conclusions

Our findings provide evidence of genetic sex differences in PD and prioritized five candidate causal proteins that appear important to sex-biased PD risk. These proteins provide insight into sex-biased PD pathobiology and therapeutic opportunities.

## Data Availability

Data used in the GWAS analyses are available upon application to:

- Blauwendraat GWAS results (https://pdgenetics.org/resources)
- FinnGen (https://www.finngen.fi/en)

Full, sex-stratified GWAS summary statistics will be available in GWAS Catalogue upon publication. The protein weights used in the study are publicly available:

- https://www.synapse.org/Synapse:syn51150434

## Authors’ contributions

N.C., Y.Z., and T.F., performed data acquisition, designed analyses, performed analyses. C.Y., S.K.S., and P.N. curated data and performed analyses. A.P.W and T.S.W. contributed data and resources. A.P.W., T.S.W., J.N.F., A.A.D., L.I., and C.C. were involved in conceptualization and study design. M.E.B. designed analyses, designed study, supervised analyses, supervised work, wrote paper, and obtained funding. All authors contributed to critical revision of the manuscript.

## Role of Funder/Sponsor

The funding organizations and sponsors had no role in the design and conduct of the study; collection, management, analysis, and interpretation of the data; preparation, review, or approval of the manuscript; and decision to submit the manuscript for publication.

## Competing interests

C.C. has received research support from GSK and EISAI. C.C. is a member of the scientific advisory board of Circular Genomics and owns stocks. C.C. is a member of the scientific advisory board of ADmit.

## Acknowledgements

This work was supported by grants from the National Institutes of Aging (R00AG075238, M.E.B). We would like to thank the following resources: IPDGC, UK Biobank, FinnGen. Detailed acknowledgements for the use of FinnGen data is provided in the supplementary material. The full FinnGen author list is provided in **eTable-11**.

