## Supplemental Document for "Sex-biased Genetic Risk Loci and Causal Brain Proteins in Parkinson’s Disease"

### Supplementary document

### Supplementary Methods

#### Genome-wide association studies

##### FinnGen

The FinnGen study is a large-scale genomics initiative that has analyzed over 500,000 Finnish biobank samples and correlated genetic variation with health data to understand disease mechanisms and predispositions. The project is a collaboration between research organisations and biobanks within Finland and international industry partners.

Ascertainment of FinnGen phenotype and genotype data is described in detail elsewhere[1]. Sex stratified analyses were conducted on the Parkinson's disease (PD) phenotype "PD_IMP" using data release R12. Documentation on Genetic data processing and statistical analyses is provided here: https://finngen.gitbook.io/documentation. FinnGen made use of Regenie to include related individuals in the genetic association analyses[2]. Information about phenotypes and endpoints is provided here: https://www.finngen.fi/en/researchers/clinical-endpoints, https://r12.risteys.finngen.fi/

#### Proteome-wide association studies

##### Brain PWAS data preparation

Human brain proteomes were previously generated from the dorsolateral prefrontal cortex (DLPFC) of donated brain samples from non-Hispanic white European (EUR) participants in the Religious Orders Study (ROS) and Memory Aging Project (MAP) cohorts[3]. Details on cohort information, genomic data QC, sample preparation, and proteomic sequencing, and data processing have been described in previous studies[4–8]. Briefly, proteomic profiling utilized isobaric tandem mass tag (TMT) peptide labelling coupled with liquid chromatography-mass spectrometry (TMT-MS). After QC, 8,458 proteins from 808 proteomic profiles were analyzed for *cis* protein quantitative trait loci (pQTLs) within ± 500 kb of the gene's boundaries (PLINK v.2.0[9]), under sex-stratified and non-sex-stratified conditions. Covariates included 56 significant surrogate variables derived from surrogate variable analysis (SVA) using the SVA package (v.3.20.0)[10] while protecting the effect of sex on protein abundance.

Non-sex-stratified and sex-stratified brain *cis* proteogenomic weights (i.e., variant protein weights) were generated using the FUSION[11] package, from prior work[6,12]. *Cis* proteogenomic weights were created in human genome build hg19/GRCh37, so PD GWAS summary statistics were thus lifted from hg38/GRCh38 to hg19/GRCh37, using the UCSC LiftOver tool[13]. PD GWAS summary statistics were intersected by genomic position and allele matching with an LD reference panel built from 1,000 Genomes Project (Phase 3)[14] EUR samples (10,871,685 variants; minor allele frequency [MAF] > 0.005). GWAS summary statistics were than processed with mungestats, filtering variants to MAF > 0.1%, and used for PWAS in FUSION[15] with default settings. Proteins with significant SNP-based heritability ($P < 0.01$) were retained. Prediction models (best linear unbiased prediction [BLUP], Bayesian sparse linear mixed model [BSLMM], least absolute shrinkage and selection operator [LASSO] regression, and elastic net [enet] regression) were evaluated, and variant protein weights were retained from the best performing model.

#### Colocalization data preparation & analysis

Variants were selected within ±1 Mb of PWAS genes' median base pair position or lead sex-stratified GWAS variants (**Methods**). Across GWAS and QTL data, non-matching variants were excluded using genomic position alignment and allele matching, while affect alleles were aligned and filtered to those displaying ≤ 10% frequency deviations across GWAS and QTL data. If respective QTL studies used smaller window sizes, then variants were inherently restricted to those overlapping PD GWAS and QTL data.

Colocalization analyses were performed as described in the main **Methods**, using coloc.abf with default per-SNP priors, which assumes a single independent signal at the locus[16], and 'coloc.susie', which incorporates SuSiE (Sum of Single Effects) regression to resolve loci with multiple independent signals[17] to validate and prioritize causal genes (coloc R package v.4.2.1)[16,17]. Coloc.abf used the p-value approach, excluding beta coefficients and standard errors to avoid potential issues with variants close to MAF 50%. Coloc.susie used the runsusie function with effect size, variance, MAF inputs, and an LD matrix from 1000 Genomes European data to identify credible sets of independent causal variants. SuSiE employs an iterative Bayesian stepwise selection method to identify credible sets of variants; if SuSiE failed to converge after 7 days runtime, the respective analysis was terminated and no results were reported. Colocalization posterior probability of hypothesis 4 (PP4) values ≥ 0.7 marked strong colocalization.

As a complementary approach, we performed colocalization analyses using ColocBoost, a recently developed method that supports both pairwise and multi-trait colocalization and is designed to better resolve shared genetic signals in regions with complex association patterns or LD structure[18]. We implemented two ColocBoost analysis strategies. First, to maintain consistency with our primary colocalization framework based on coloc.abf and coloc.susie, we performed pairwise ColocBoost analyses between each sex-stratified PD GWAS and the corresponding discovery pQTL dataset (either the sex-matched pQTL dataset for primary discovery or the non-sex-stratified pQTL dataset for secondary discovery). Second, we performed multi-trait ColocBoost analyses that jointly modeled the sex-stratified PD GWAS, the sex-matched pQTL dataset, and the non-sex-stratified pQTL dataset, leveraging the multi-trait capabilities uniquely supported by ColocBoost.

For each sex-biased PWAS signal, we defined a ±1 Mb window around the gene midpoint and extracted summary statistics for all variants within the region. Z-scores were obtained for the sex-stratified PD GWAS and all relevant pQTL datasets. pQTL effect alleles were then harmonized to the GWAS reference and alternative allele coding, and analyses were restricted to variants present across all datasets included in a given ColocBoost model. An LD correlation matrix for the analyzed variants was computed from European samples in the 1000 Genomes Project using PLINK2[9]. The colocboost function was then run using the corresponding Z-scores, sample sizes, and LD matrix, with the PD GWAS specified as the focal outcome and all other parameters set to their default values. For pairwise analyses, the model included the PD GWAS and a single pQTL dataset. For multi-trait analyses, the model jointly included the PD GWAS, sex-matched pQTL dataset, and non-sex-stratified pQTL dataset. For each locus, the resulting cos_summary output was retained, and the cos_npc metric was extracted as a measure analogous to the posterior probability of colocalization (PP4) reported by coloc.abf and coloc.susie.

#### SMR/HEIDI analysis

The Summary-based Mendelian Randomization (SMR) method is a statistical framework used to test for pleiotropic or potentially causal associations between gene expression (or other molecular traits) and complex diseases or phenotypes by integrating summary-level data from genome-wide association studies (GWAS) with molecular quantitative trait loci (xQTL) datasets, such as expression QTL (eQTL), DNA methylation QTL (mQTL), and protein QTL (pQTL)[19]. In our SMR analysis, the top associated cis-pQTL was selected as an instrumental variable to evaluate whether variation in the protein level is associated with the risk of PD. For each candidate gene identified by PWAS, GWAS summary statistics were restricted to the corresponding cis region, and pQTL datasets were converted into BESD format following the pipeline provided by Yang Lab[19]. SMR tests were considered supportive at FDR-P<0.05, but additionally required a negative finding on the HEIDI test. The HEIDI (Heterogeneity In Dependent Instruments) test was applied to distinguish true causal or pleiotropic associations from those driven by linkage disequilibrium (LD). Only SNPs with P-values below $1.57 \times 10^{-3}$ were included in the HEIDI test. These SNPs were further pruned based on LD, retaining variants with $r^2$ between 0.05 and 0.9. A minimum of three and a maximum of twenty cis-SNPs were used for each HEIDI test. Associations with non-significant HEIDI results (P>0.05) were considered consistent with a shared causal variant, whereas significant HEIDI results (P<0.05) indicated potential confounding due to LD, and such SMR results were considered not supported. All analyses were conducted using the SMR software (version 1.3.1).

#### Locus Novelty

For our GWAS lead variants, we queried the Open Target database[20] to assess if lead variants are in any GWAS credible sets reported in a Parkinson's disease phenotype. If found, we reported the gene linked to the lead variants of the credible set. If no match was found, we consulted two additional studies, Kim et al.[21] and Nalls et al.[22], and assigned the reported gene to our lead variant if it fell within 1 Mb of a reported variant and had an LD $R^2 > 0.1$. Variants that met none of these criteria were annotated as novel.

### Acknowledgements

#### Acknowledgments for other GWAS and phenotype data

##### *FinnGen Study*

We want to acknowledge the participants and investigators of the FinnGen study. The FinnGen project is funded by two grants from Business Finland (HUS 4685/31/2016 and UH 4386/31/2016) and the following industry partners: AbbVie Inc., Alnylam Pharmaceuticals, Inc., AstraZeneca UK Ltd, Bayer AG, Biogen MA Inc., Boehringer Ingelheim International GmbH, Bristol Myers Squibb Inc. (and Celgene Corporation & Celgene International II Sàrl), Genentech Inc., GlaxoSmithKline Intellectual Property Development Ltd., Johnson&Johnson Innovative Medicine Inc., Maze Therapeutics Inc., Merck Sharp & Dohme LCC, Novartis AG, Pfizer Inc. and Sanofi US Services Inc. Following biobanks are acknowledged for delivering biobank samples to FinnGen: Auria Biobank (www.auria.fi/biopankki), THL Biobank (www.thl.fi/biobank), Helsinki Biobank (www.helsinginbiopankki.fi), Biobank Borealis of Northern Finland (https://www.ppshp.fi/Tutkimus-ja-opetus/Biopankki/Pages/Biobank-Borealis-briefly-in-English.aspx), Finnish Clinical Biobank Tampere (www.tays.fi/en-US/Research_and_development/Finnish_Clinical_Biobank_Tampere), Biobank of Eastern Finland (www.ita-suomenbiopankki.fi/en), Central Finland Biobank (www.ksshp.fi/fi-FI/Potilaalle/Biopankki), Finnish Red Cross Blood Service Biobank (www.veripalvelu.fi/verenluovutus/biopankkitoiminta), Terveystalo Biobank (www.terveystalo.com/fi/Yritystietoa/Terveystalo-Biopankki/Biopankki/) and Arctic Biobank (https://www.oulu.fi/en/university/faculties-and-units/faculty-medicine/northern-finland-birth-cohorts-and-arctic-biobank). All Finnish Biobanks are members of BBMRI.fi infrastructure (https://www.bbmri-eric.eu/national-nodes/finland/). Finnish Biobank Cooperative -FINBB (https://finbb.fi/) is the coordinator of BBMRI-ERIC operations in Finland. The Finnish biobank data can be accessed through the Fingenious® services (https://site.fingenious.fi/en/) managed by FINBB.

##### FinnGen ethics statement

Study subjects in FinnGen provided informed consent for biobank research, based on the Finnish Biobank Act. Alternatively, separate research cohorts, collected prior the Finnish Biobank Act came into effect (in September 2013) and start of FinnGen (August 2017), were collected based on study-specific consents and later transferred to the Finnish biobanks after approval by Fimea (Finnish Medicines Agency), the National Supervisory Authority for Welfare and Health. Recruitment protocols followed the biobank protocols approved by Fimea. The Coordinating

Ethics Committee of the Hospital District of Helsinki and Uusimaa (HUS) statement number for the FinnGen study is Nr HUS/990/2017.

The FinnGen study is approved by Finnish Institute for Health and Welfare (permit numbers: THL/2031/6.02.00/2017, THL/1101/5.05.00/2017, THL/341/6.02.00/2018, THL/2222/6.02.00/2018, THL/283/6.02.00/2019, THL/1721/5.05.00/2019 and THL/1524/5.05.00/2020), Digital and population data service agency (permit numbers: VRK43431/2017-3, VRK/6909/2018-3, VRK/4415/2019-3), the Social Insurance Institution (permit numbers: KELA 58/522/2017, KELA 131/522/2018, KELA 70/522/2019, KELA 98/522/2019, KELA 134/522/2019, KELA 138/522/2019, KELA 2/522/2020, KELA 16/522/2020), Findata permit numbers THL/2364/14.02/2020, THL/4055/14.06.00/2020, THL/3433/14.06.00/2020, THL/4432/14.06/2020, THL/5189/14.06/2020, THL/5894/14.06.00/2020, THL/6619/14.06.00/2020, THL/209/14.06.00/2021, THL/688/14.06.00/2021, THL/1284/14.06.00/2021, THL/1965/14.06.00/2021, THL/5546/14.02.00/2020, THL/2658/14.06.00/2021, THL/4235/14.06.00/2021, Statistics Finland (permit numbers: TK-53-1041-17 and TK/143/07.03.00/2020 (earlier TK-53-90-20) TK/1735/07.03.00/2021, TK/3112/07.03.00/2021) and Finnish Registry for Kidney Diseases permission/extract from the meeting minutes on 4th July 2019.

The Biobank Access Decisions for FinnGen samples and data utilized in FinnGen Data Freeze 12 include: THL Biobank BB2017_55, BB2017_111, BB2018_19, BB_2018_34, BB_2018_67, BB2018_71, BB2019_7, BB2019_8, BB2019_26, BB2020_1, BB2021_65, Finnish Red Cross Blood Service Biobank 7.12.2017, Helsinki Biobank HUS/359/2017, HUS/248/2020, HUS/430/2021 §28, §29, HUS/150/2022 §12, §13, §14, §15, §16, §17, §18, §23, §58, §59, HUS/128/2023 §18, Auria Biobank AB17-5154 and amendment #1 (August 17 2020) and amendments BB_2021-0140, BB_2021-0156 (August 26 2021, Feb 2 2022), BB_2021-0169, BB_2021-0179, BB_2021-0161, AB20-5926 and amendment #1 (April 23 2020) and it´s modifications (Sep 22 2021), BB_2022-0262, BB_2022-0256, Biobank Borealis of Northern Finland_2017_1013, 2021_5010, 2021_5010 Amendment, 2021_5018, 2021_5018 Amendment, 2021_5015, 2021_5015 Amendment, 2021_5015 Amendment_2, 2021_5023, 2021_5023 Amendment, 2021_5023 Amendment_2, 2021_5017, 2021_5017 Amendment, 2022_6001, 2022_6001 Amendment, 2022_6006 Amendment, 2022_6006 Amendment, 2022_6006 Amendment_2, BB22-0067, 2022_0262, 2022_0262 Amendment, Biobank of Eastern Finland 1186/2018 and amendment 22§/2020, 53§/2021, 13§/2022, 14§/2022, 15§/2022, 27§/2022, 28§/2022, 29§/2022, 33§/2022, 35§/2022, 36§/2022, 37§/2022, 39§/2022, 7§/2023, 32§/2023, 33§/2023, 34§/2023, 35§/2023, 36§/2023, 37§/2023, 38§/2023, 39§/2023, 40§/2023, 41§/2023, Finnish Clinical Biobank Tampere MH0004 and amendments (21.02.2020 & 06.10.2020), BB2021-0140 8§/2021, 9§/2021, §9/2022, §10/2022, §12/2022, 13§/2022, §20/2022, §21/2022, §22/2022, §23/2022, 28§/2022, 29§/2022, 30§/2022, 31§/2022, 32§/2022, 38§/2022, 40§/2022, 42§/2022, 1§/2023, Central Finland Biobank 1-2017, BB_2021-0161, BB_2021-0169, BB_2021-0179, BB_2021-0170, BB_2022-0256, BB_2022-0262, BB22-0067, Decision allowing to continue data processing until 31st Aug 2024 for projects: BB_2021-0179, BB22-0067,BB_2022-0262, BB_2021-0170, BB_2021-0164, BB_2021-0161, and BB_2021-0169, and Terveystalo Biobank STB 2018001 and amendment 25th Aug 2020, Finnish Hematological Registry and Clinical Biobank decision 18th June 2021, Arctic biobank P0844: ARC_2021_1001.

### Supplementary Figures

#### GWAS

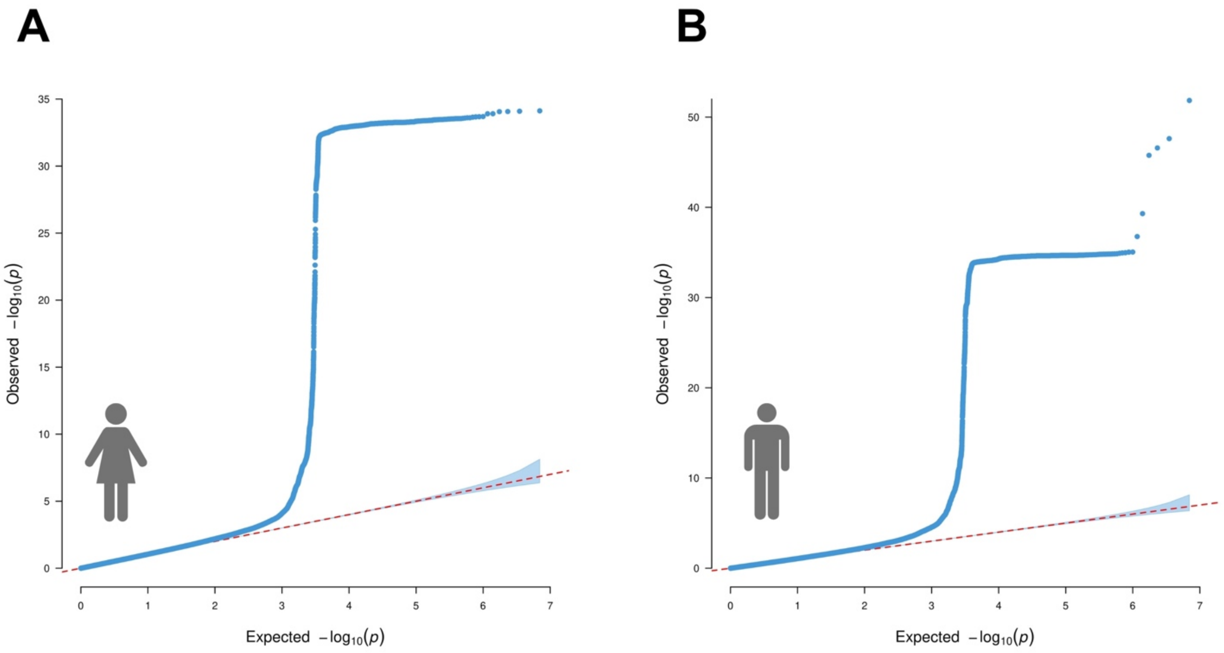

**Figure S1. Quantile-Quantile (QQ) plots corresponding to sex-stratified GWAS of Parkinson's disease. A)** Women. The inflation factor ($\lambda$=1.0525) and sample size-adjusted inflation factor ($\lambda_{1,000}$=1.0001) showed no sign of inflation. **B)** Men. The inflation factor ($\lambda$=1.0711) and sample size-adjusted inflation factor ($\lambda_{1,000}$=1.0002) showed no sign of inflation.

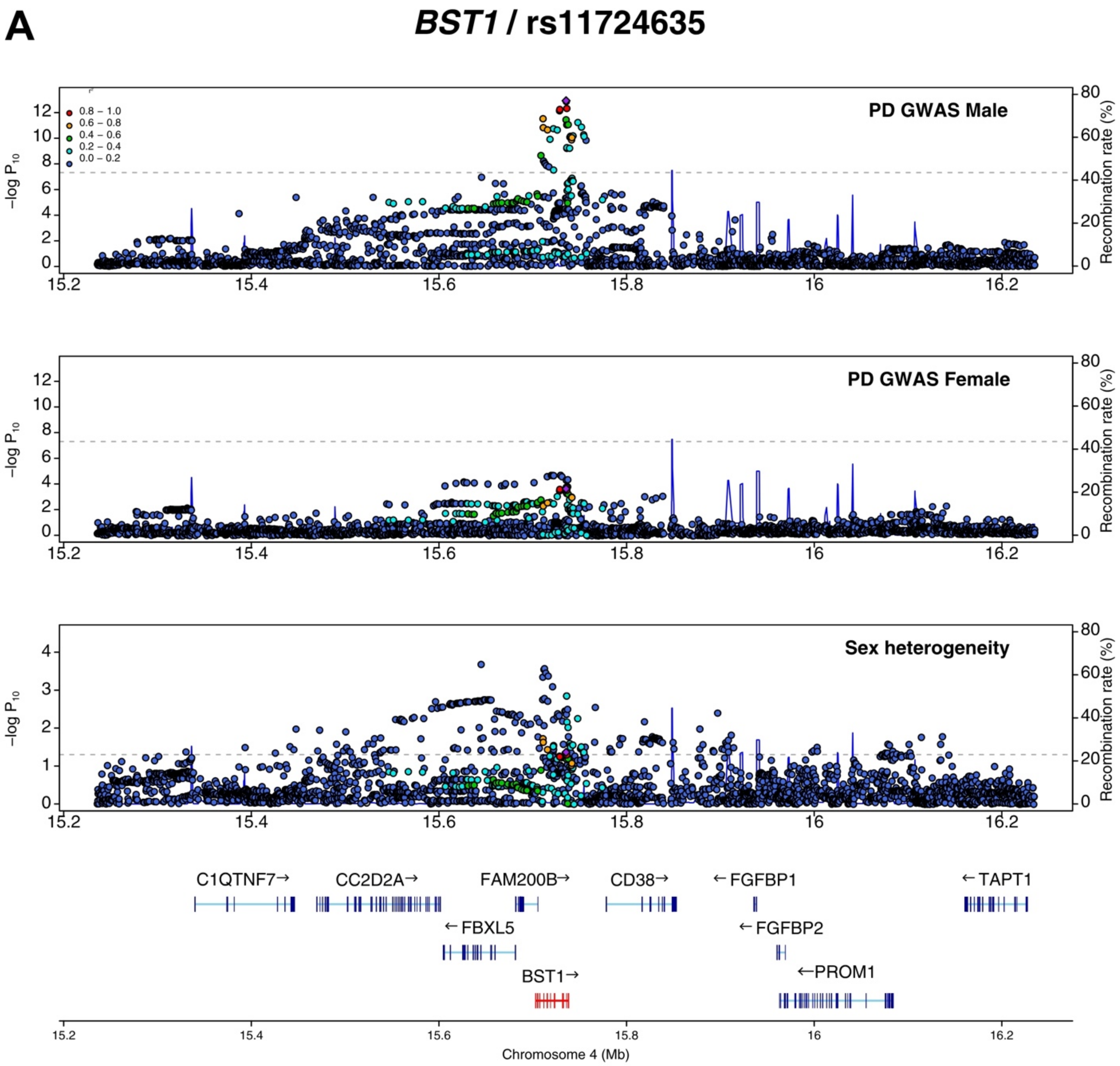
A
BST1 / rs11724635
PD GWAS Male
PD GWAS Female
Sex heterogeneity
0.8 – 1.0
0.6 – 0.8
0.4 – 0.6
0.2 – 0.4
0.0 – 0.2
−log P10
Recombination rate (%)
C1QTNF7→
CC2D2A→
FAM200B→
CD38→
←FGFBP1
←TAPT1
←FBXL5
←FGFBP2
BST1→
←PROM1
Chromosome 4 (Mb)

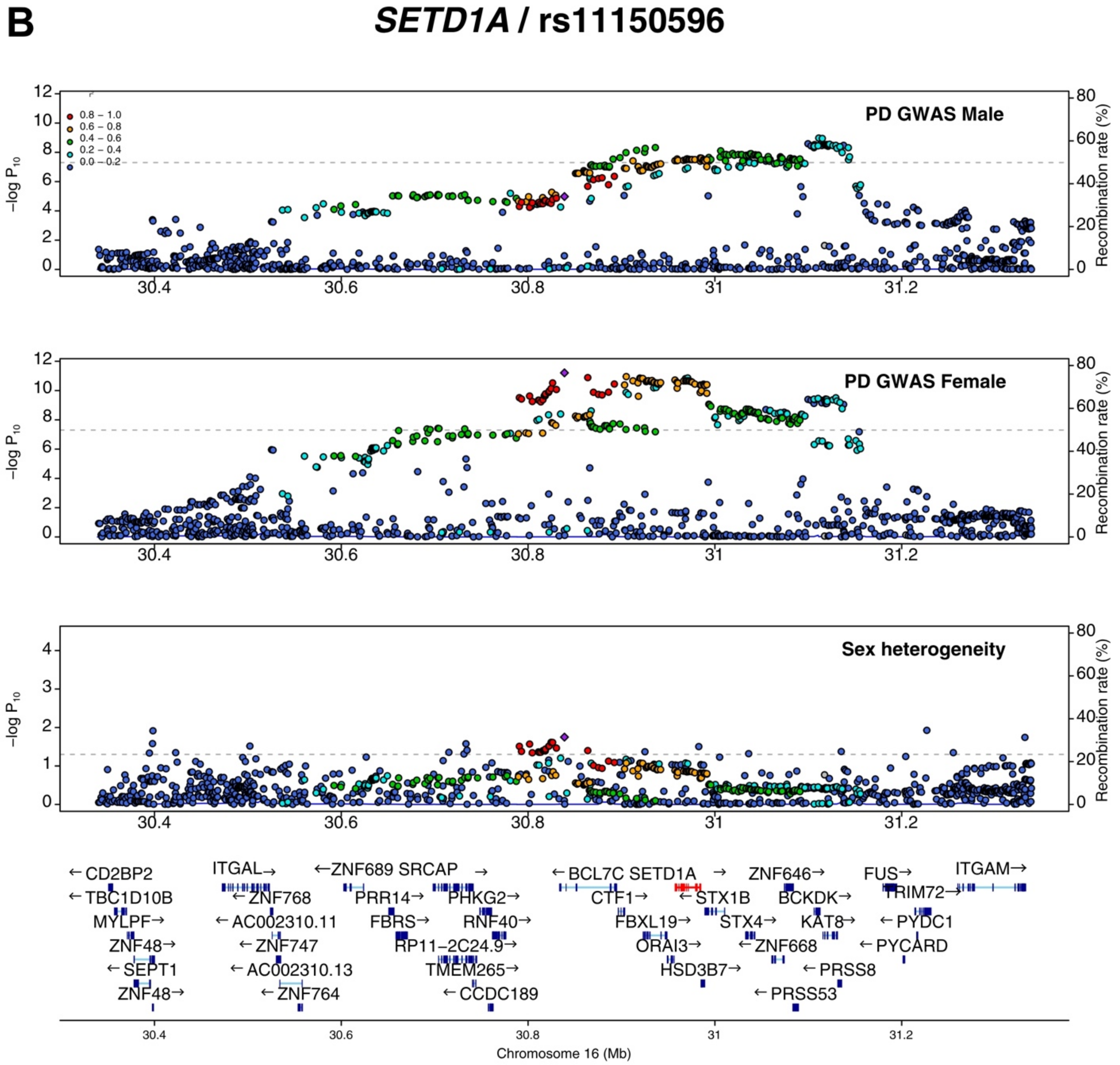

**Figure S2. Locus zoom plot of nominal sex-heterogenous Parkinson's disease GWAS lead variants. A)** *BST1*; **B)** *SETD1A*. Dot color coding represents linkage with tagging variant (purple diamond): red - R2: 0.8-1.0, orange - R2:0.6-0.8, green - R2:0.4-0.6, cyan - R2:0.2-0.4, blue - R2:0.0-0.2.

#### PWAS

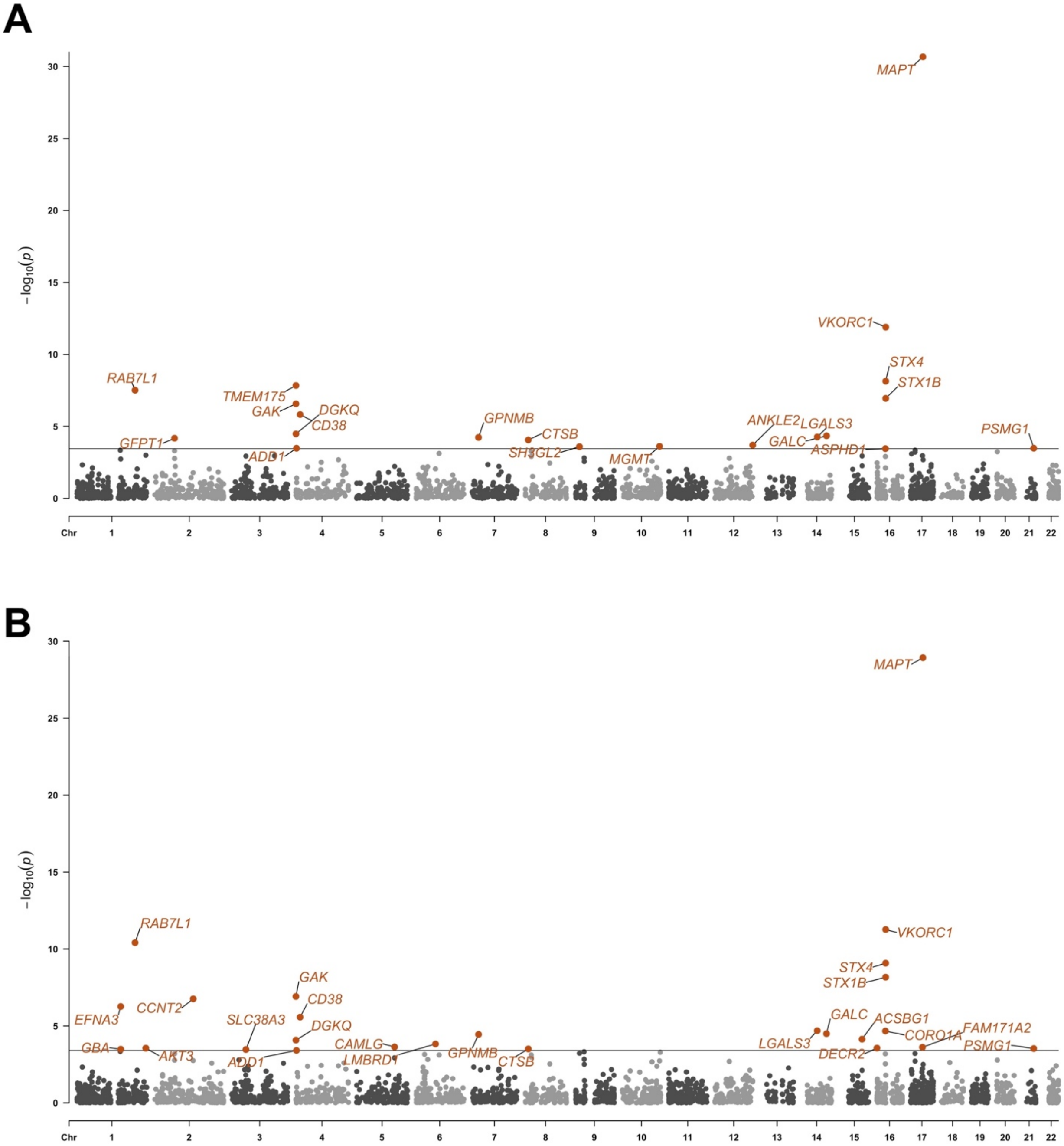
A
MAPT
VKORC1
STX4
STX1B
RAB7L1
TMEM175
GAK
DGKQ
CD38
GFPT1
ADD1
GPNMB
CTSB
SH3GL2
MGMT
ANKLE2
LGALS3
GALC
ASPHD1
PSMG1
B
MAPT
RAB7L1
VKORC1
STX4
STX1B
GAK
CD38
CCNT2
EFNA3
SLC38A3
DGKQ
GALC
ACSBG1
FAM171A2
LGALS3
CORO1A
PSMG1
DECR2
GBA
AKT3
CAMLG
LMBRD1
GPNMB
CTSB
ADD1
$-\log_{10}(p)$
Chr 1 2 3 4 5 6 7 8 9 10 11 12 13 14 15 16 17 18 19 20 21 22

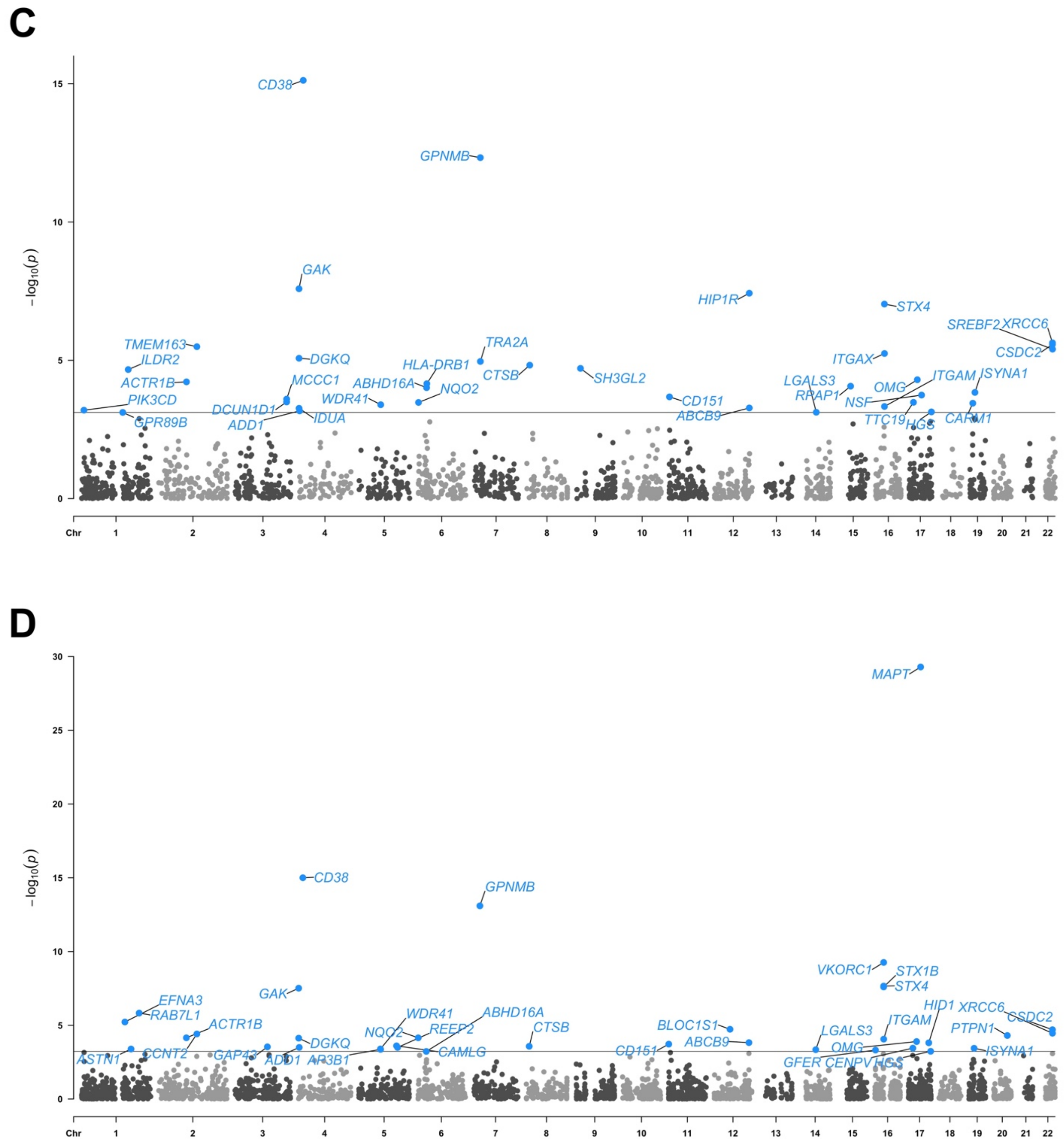

**Figure S3. Sex-stratified Parkinson's disease PWAS: All discoveries by sex and design. A)** Female primary discovery; **B)** Female secondary discovery; **C)** Male primary discovery; **D)** Male secondary discovery. Datapoints in the Manhattan plots represent association tests between PD and a given protein in brain proteogenomic data. Primary discoveries integrate the sex-stratified GWAS with sex-matched protein weights, while secondary discoveries integrate the sex-stratified GWAS with non-sex-stratified protein weights. X-axis denotes genomic positions, and y-axis represents -log10($P$) from PWAS findings; the solid gray horizontal line denotes the proteome-wide significance threshold at $P_{FDR}$<0.05.

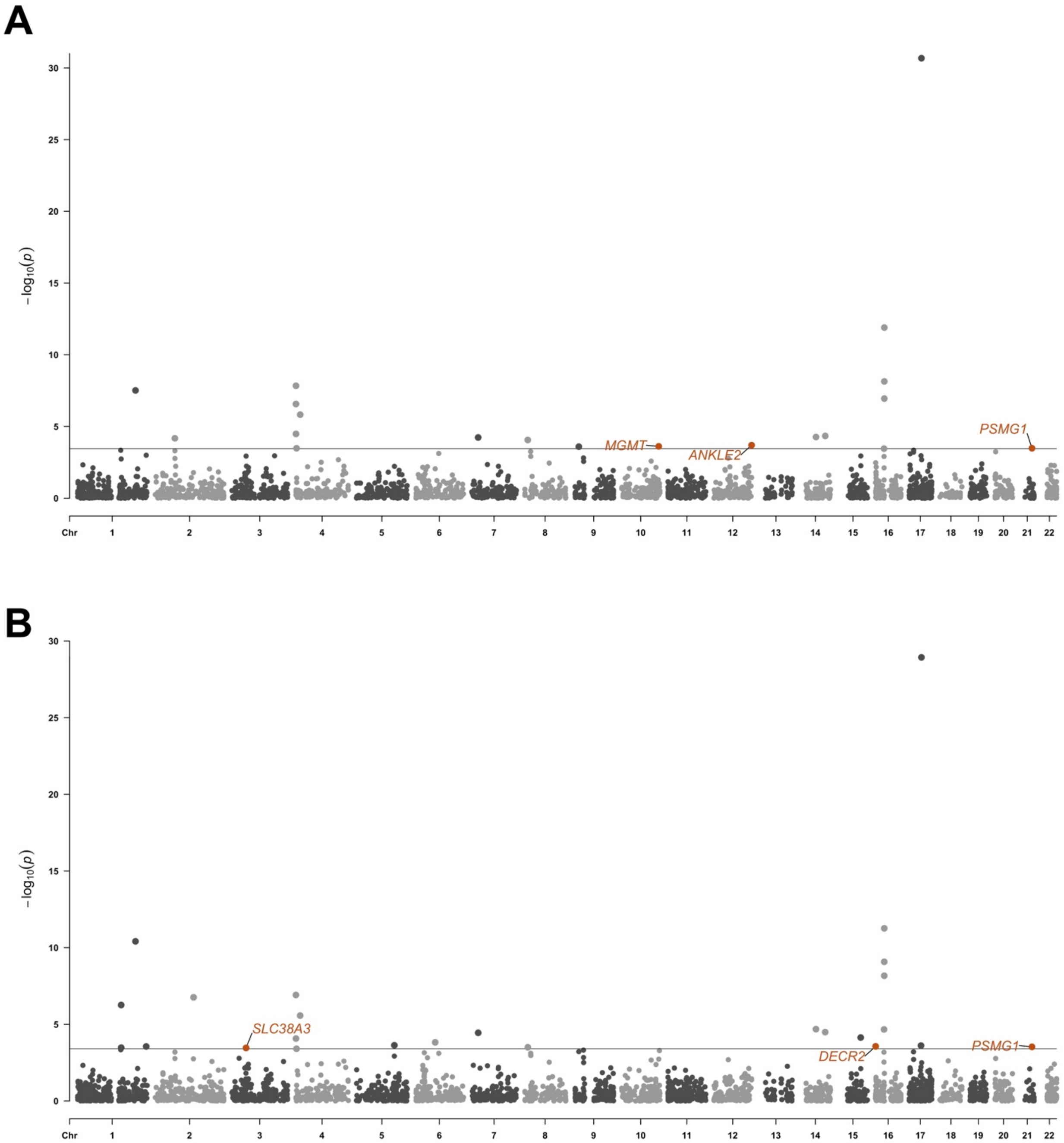

A
B
$-\log_{10}(p)$
Chr
MGMT
ANKLE2
PSMG1
SLC38A3
DECR2
PSMG1

C

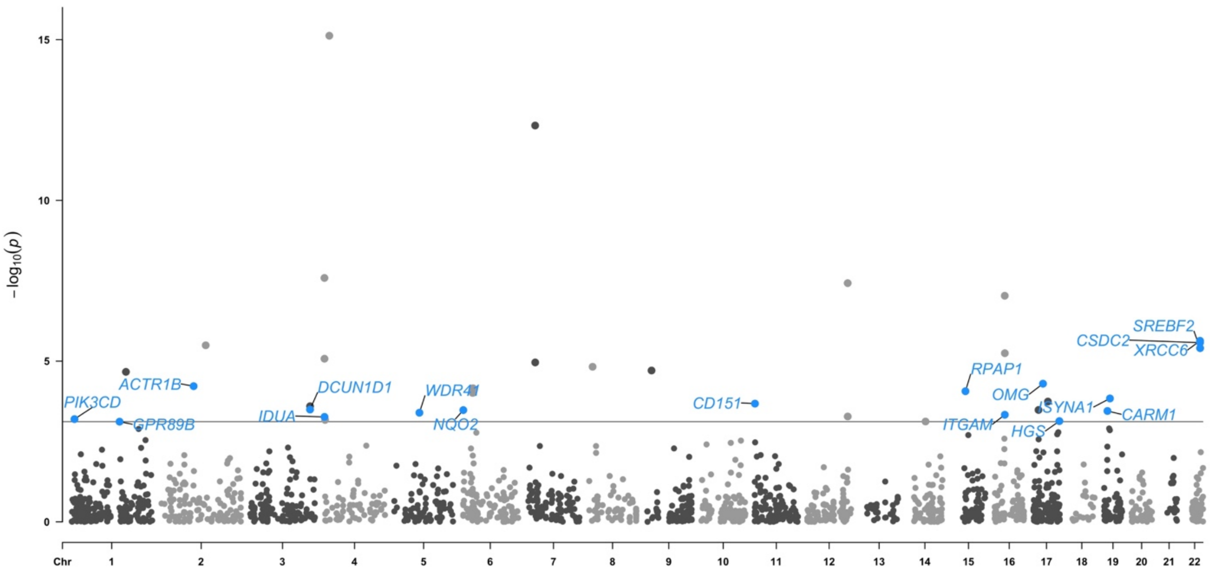

D

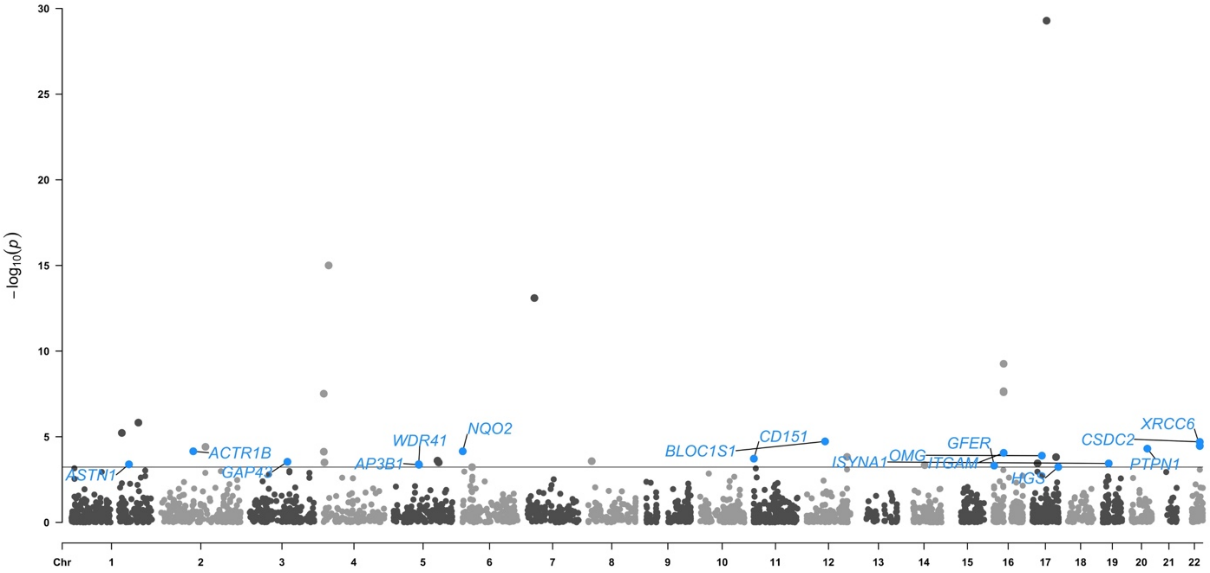

**Figure S4. Sex-stratified Parkinson's disease PWAS: Sex-specific discoveries by sex and design. A)** Female primary discovery; **B)** Female secondary discovery; **C)** Male primary discovery; **D)** Male brain discovery. Datapoints in the Manhattan plots represent association tests between PD and a given protein in brain proteogenomic data. Only genes passing sex-heterogeneity filters are annotated and colored based on which sex the gene was prioritized in. X-axis denotes genomic positions, and y-axis represents -log10(*P*) from PWAS analyses; the solid gray horizontal line denotes the proteome-wide significance threshold at $P_{FDR}$<0.05.

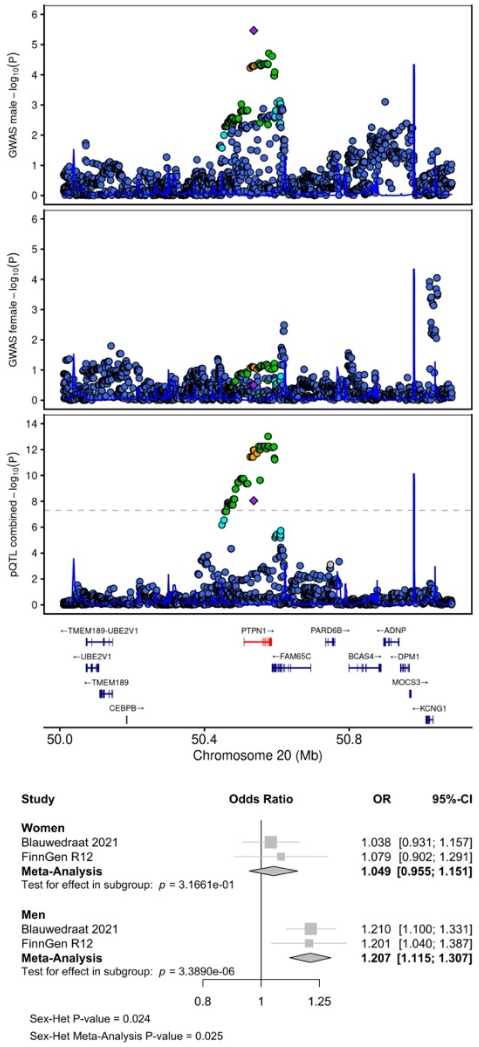

**Figure S5. Locus zoom and forest plots for PTPN1.** Locus zoom plots for male PD GWAS (top), female PD GWAS (middle), and non-sex-matched colocalized pQTL data (bottom). The same tagging variant is indicated throughout**.** Dot color coding represents linkage with tagging variant (purple diamond): red - R2: 0.8-1.0, orange - R2:0.6-0.8, green - R2:0.4-0.6, cyan - R2:0.2-0.4, blue - R2:0.0-0.2. Forest plots indicate PD associations across cohorts and sexes for the tagging variant.
